# Analytical approaches for antimalarial antibody responses to confirm historical and recent malaria transmission: an example from the Philippines

**DOI:** 10.1101/2022.06.16.22276488

**Authors:** Maria Lourdes M. Macalinao, Kimberly M. Fornace, Ralph A. Reyes, Alison Paolo N. Bareng, Tom Hall, John H. Adams, Christèle Huon, Chetan E. Chitnis, Jennifer S. Luchavez, Kevin K. A. Tetteh, Katsuyuki Yui, Julius Clemence R. Hafalla, Fe Esperanza J. Espino, Chris J. Drakeley

## Abstract

**Background:** Assessing the status of malaria transmission in endemic areas becomes increasingly challenging as countries approach elimination. Serology can provide robust estimates of malaria transmission intensities, and multiplex serological assays allow for simultaneous assessment of markers of recent and historical malaria exposure.

**Methods:** Here, we evaluated different statistical and machine learning methods for analyzing multiplex malaria-specific antibody response data to classify recent and historical exposure to *Plasmodium falciparum* and *P. vivax*. To assess these methods, we utilized samples from a health-facility based survey (n=9132) in the Philippines, where we quantified antibody responses against 8 *P. falciparum* and 6 *P. vivax*-specific antigens from 3 sites with varying transmission intensity.

**Findings:** Measurements of antibody responses and seroprevalence were consistent with the 3 sites’ known endemicity status. For predicting *P. falciparum* infection, a machine learning (ML) approach (Random Forest model) using 4 serological markers (PfGLURP R2, Etramp5.Ag1, GEXP18 and PfMSP1_19_) gave better predictions for cases in Palawan (AUC: 0·9591, CI 0·9497-0·9684) than individual antigen seropositivity. Although the ML approach did not improve *P. vivax* infection predictions, ML classifications confirmed the absence of recent exposure to *P. falciparum* and *P. vivax* in both Occidental Mindoro and Bataan. For predicting historical *P. falciparum* and *P. vivax* transmission, seroprevalence and seroconversion rates based on cumulative exposure markers AMA1 and MSP1_19_ showed reliable trends in the 3 sites.

**Interpretation:** Our study emphasizes the utility of serological markers in predicting recent and historical exposure in a sub-national elimination setting, and also highlights the potential use of machine learning models using multiplex antibody responses to improve assessment of the malaria transmission status of countries aiming for elimination. This work also provides baseline antibody data for monitoring risk in malaria-endemic areas in the Philippines.

**Funding:** Newton Fund, Philippine Council for Health Research and Development, and UK Medical Research Council.

## Introduction

In malaria elimination areas, where reporting of local cases continually decline, assessing and differentiating areas with residual transmission becomes increasingly challenging. In the case of the Philippines, which is aiming to eliminate malaria by 2030 following a sub-national elimination approach^1^, the country saw >70% decrease in malaria cases in the past decade, with only 2 of 81 provinces currently reporting local cases in 2021, and 19 in elimination phase. *Plasmodium falciparum* (Pf) contributes > 80% to the total malaria cases, *P. vivax* (Pv) is at >20%, while the other species *P. malariae, P. ovale* and *P. knowlesi* make up <1% of the cases^2^. Innovative tools that are capable of detecting both past and present infections can potentially be used to confirm the presence or absence of malaria transmission in this kind of setting that employs subnational specific control approaches^3–6^.

Several studies have utilized serology and malaria-specific antibody responses to estimate malaria transmission intensities^7,8^, showing that these represent a viable additional metric of both historical and recent exposure^9–14^. Antibody prevalence alone and as age-adjusted seroconversion rates correlate with entomological and parasitological measures used in estimating malaria transmission^15–17^. Many of the original studies examined single antigen platforms and antigens associated with cumulative exposure to infection such as Pf apical membrane antigen-1 (PfAMA1) and the 19KDa fragment of Pf merozoite protein 1 (PfMSP1_19_). More recent advances in array and bead-based assay platforms allow for simultaneous analysis of antibody responses to multiple antigens^12,18–22^. These approaches allow the inclusion of multiple targets that can represent diversity in the parasite and variation in individual immune response. Recent multi-antigen studies have identified markers associated with antibody responses describing recent and historical *P. falciparum* and *P. vivax* exposure^13,23–25^. To fully realize the additional information provided by examining multiple antigenic targets, more advanced statistical approaches and algorithms such as machine learning can be employed to predict optimal combinations of antibody responses for the outcome of interest.

The overall aim of this study was to evaluate known malaria-specific *P. falciparum* and *P. vivax* serological markers for their predictive capacity to distinguish current or recent infections from historically exposed individuals. Specifically, we sought to evaluate different approaches to determining seropositivity for estimating malaria transmission intensities and exposure in areas of varying endemicity, and apply this using serological data from the Philippines. To achieve this, we: 1) evaluated analysis methods to determine seropositivity using malaria-specific antibody responses to single and multiple antigens, utilizing multiplex serological data from health facility surveys in 3 sites in the Philippines, and 2) analyzed antibody responses and estimated transmission intensities in relation to the supposed immune status of these populations. Our findings detailing the antibody responses to multiple malaria-specific antigens demonstrate the utility of serology in showing the heterogeneity of malaria transmission in malaria-endemic populations in the Philippine setting.

## Methods

### Ethical approval

This study was reviewed and approved by the Research Institute for Tropical Medicine – Institutional Review Board (RITM IRB 2016-04) and LSHTM Research Ethics Committee (11597).

### Data Source: Study Sites and Samples

The study was conducted in 3 municipalities in 3 provinces in the Philippines, representing areas of varying malaria endemicity: Rizal, Palawan, currently the most endemic municipality in the Philippines, and reported >60% of the total cases in the country (annual parasite index, API of 5·7 per 1,000 risk population) in 2018; Abra de Ilog, Occidental Mindoro, a municipality reporting sporadic local cases and with declining transmission (API of 0·38 in 2018); and Morong, Bataan with a last reported indigenous case in 2011 and declared malaria-free in 2019 (Figure 1).

**Figure 1.**
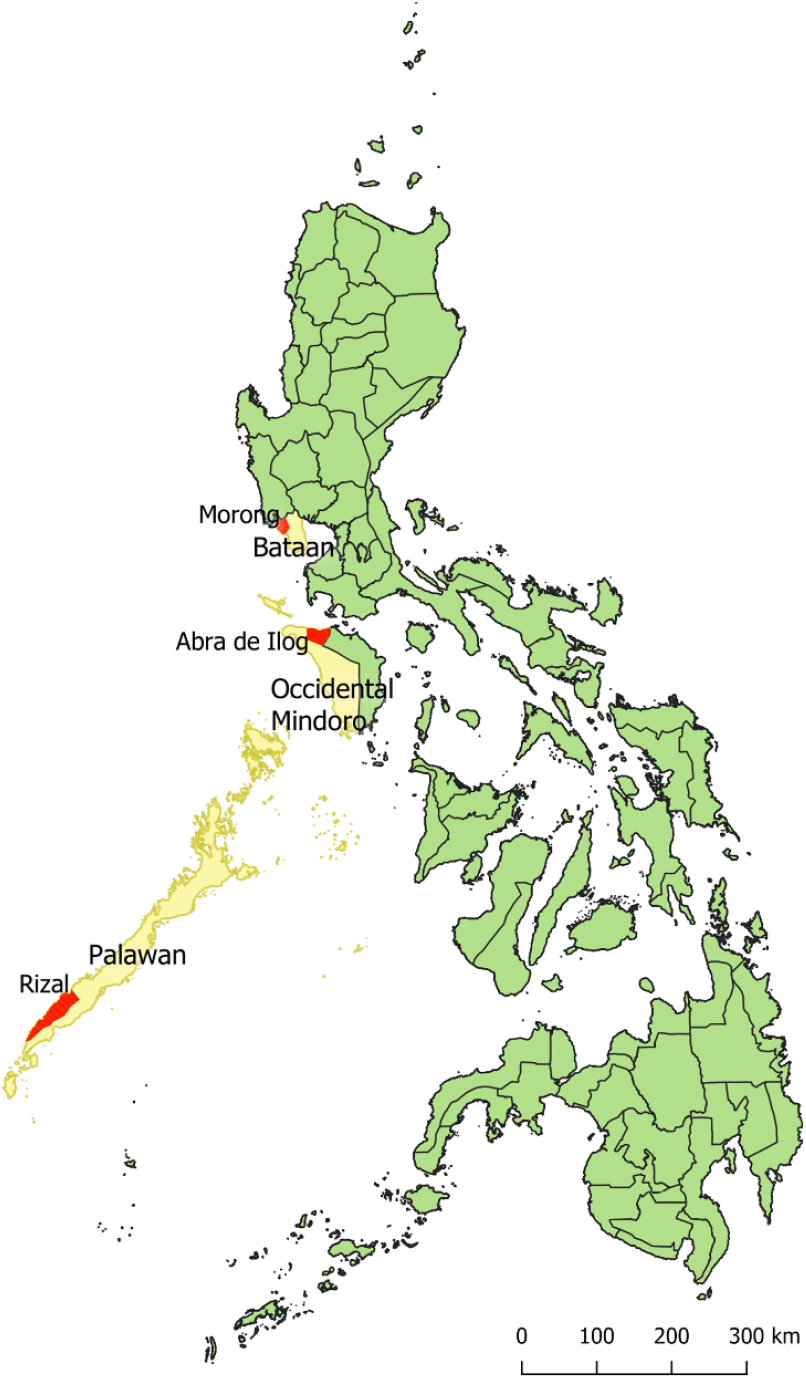
Map showing the study sites, with red areas as the focused municipalities within the provinces marked yellow.

Participants were recruited from June 2016 to June 2018 in a health facility-based rolling cross-sectional survey detailed in Reyes et al **(2021)**. Briefly, all consulting patients, as well as their companions, were invited to participate and provide a finger-pricked blood sample for malaria diagnosis through microscopy (blood smear), rapid diagnostic test (RDT), and a dried blood spot sample (DBS) on Whatman 3MM CHR filter paper for malaria diagnosis by PCR and serological analysis.

### Multiplex bead-based assay of malaria-specific antibodies

Serological analysis was conducted using a multiplex bead-based assay as previously described^23,27,28^, with an antigen panel that included 8 *P. falciparum*-specific and 6 *P. vivax*-specific recombinant antigens coupled to Magplex beads (Luminex Corp, Austin, TX, USA). The antigens were PfAMA1 (apical membrane antigen 1), PfMSP1_19_ (merozoite surface protein), and their *P. vivax* homologues PvAMA1 and PvMSP1_19_; PfGLURP R2 (glutamate rich protein), Etramp5.Ag1 (early transcribed membrane protein 5), PfSEA1 (schizont egress antigen), GEXP18 (Gametocyte exported protein 18), MSP2 CH150/9 (CH150/9 allele of MSP2), MSP 2 Dd2 (Dd2 allele of MSP2), PvEBP (erythrocyte binding protein), PvRBP1a (reticulocyte binding protein 1a), PvRII and PvDBPRII (region II, Duffy binding protein). Antigen characteristics are shown in Table 1.

**Table 1.** List of antigens in the multiplex bead-based assay panel

| Antigen | Description | Gene ID | Reference |
| --- | --- | --- | --- |
| <u>Pf-specific</u> |  |  |  |
| PfAMA1 | apical membrane antigen 1 | PF3D7_1133400 | Collins et al., 2007 |
| PfMSP1<sub>19</sub> | 19 kDa fragment of merozoite surface protein (MSP) 1 | PF3D7_0930300 | Burghaus and Holder, 1994 |
| PfGlurp R2 | glutamate rich protein R2 | PF3D7_1035300 | Theisen et al., 1995 |
| Etramp5.Ag1 | early transcribed membrane protein 5 | PF3D7_0532100 | van den Hoogen et al., 2019; Spielmann et al., 2003 |
| PfSEA1 | schizont egress antigen | PF3D7_1021800 | Raj et al., 2014 |
| GEXP18 | Gametocyte exported protein 18 | PF3D7_0402400 | Helb et al., 2015 |
| MSP2 CH150 | CH150/9 allele of MSP2 | PF3D7_0206800 | Polley et al., 2006 |
| MSP2 Dd2 | Dd2 allele of MSP2 | PF3D7_0206800 | Taylor et al., 1995 |
| <u>Pv-specific</u> |  |  |  |
| PvAMA1 | apical membrane antigen 1 | PVX_092275 | Chuquiyauri et al., 2015 |
| PvMSP1<sub>19</sub> | merozoite surface protein | PVX_099980 | França et al., 2016b |
| PvRII | region II, Duffy binding protein | PVX_110810 | França et al., 2016a |
| PvDBP RII | region II, Duffy binding protein | PVX_110810 | Ntumngia et al., 2012 |
| PvEBP | erythrocyte binding protein | PVX_110835 | Hester et al., 2013; Menard et al., 2013 |
| PvRBP 1a | reticulocyte binding protein 1a | PVX_098585 | França et al., 2016a |

The assay was conducted as described in Wu et al (**2019**). Briefly, serum was eluted from DBS samples (∼1uL from 3mm punch; 1:400 final dilution). Approximately 1000 beads per antigen were added per well in 96-well flat bottom plates, which are incubated for 1·5 hours on a shaker with 50uL sample and controls (2 positive controls for *P. falciparum* and *P. vivax* – pooled plasma from adults in hyperendemic malaria setting; 1 negative control – European malaria-naïve blood donors). After washing, samples are incubated for 1·5 hours with 50uL 1:200 goat anti-human Fcy-fragment-specific IgG (Jackson Immuno 109-116-098: conjugated to R-PE). Samples were washed, suspended in PBS, and read using a Luminex 200 or Magpix machine, with net median fluorescence intensity (MFI) levels to all antigens recorded for all samples. Plate specific adjustments were performed based on the outcome of standard control curves generated from positive control pools included on each plate.

### Data analysis

Statistical analyses were performed using R version 3·6·3 and Graphpad Prism 8. IgG antibody responses recorded as net MFI values were analyzed using different methods. Quantitative continuous antibody response data (reported as log_10_ MFI values) were compared for different groups (*i*.*e*., by age group, study site or current malaria infection) using Student’s t-test or one-way ANOVA Kruskal-Wallis test with Wilcoxon test for pairwise comparisons. Correlations between the levels of antibody responses as well as age and malaria positivity were analyzed using Spearman’s rank correlation.

### Determining seropositivity rates using single antigen responses

Binary outcomes for seropositivity to each antigen were determined through the computation of cut-off values in 2 ways: 1) a 2-component finite mixture model (FMM); and 2) a reference negative population (NegPop) model. FMM was computed using the mixEM function in R mixtools package. For the NegPop model, European naïve samples were used as reference population. For both models, samples were considered seropositive for specific antigens if MFI values are higher than the antigen cutoff values (mean MFI plus 3 standard deviations). To evaluate the accuracy of these classifications, we used samples from known exposed and unexposed populations as validation data (Table 2), comprising *P. falciparum* (Pf+) and *P. vivax*-positive (Pv+) datasets, and a malaria-negative (neg) dataset collated from previous studies and this study. Sensitivity and specificity for identifying current *P. falciparum* and *P. vivax* infection from this study and the validation dataset, and receiver operating characteristics (ROC) curves, were determined for each serological marker (Supplementary Tables 1 and 2).

**Table 2.** Training and validation data used in the classification models

| Training data used | N | Validation dataset * | Negative population model | SuperLearner / Random Forest: Recent Pf exposure | SuperLearner / Random Forest: Recent Pv exposure | SuperLearner / Random Forest: Historical Pf exposure | Random Forest: Historical Pf exposure | Random Forest: Historical Pv exposure |
| --- | --- | --- | --- | --- | --- | --- | --- | --- |
| **European naive controls** | 179 | **neg** | — | — | — | — | — | — |
| **Positive controls** |  |  |  |  |  |  |  |  |
| African *P. falciparum*-positive | 4 | **Pf+** |  | + |  |  |  |  |
| African *P. vivax*-positive | 10 | **Pv+** |  |  | + |  |  |  |
| **This study** |  |  |  |  |  |  |  |  |
| *P. falciparum*-positive subset | 568 | **Pf+** |  | + |  |  |  |  |
| *P. vivax* microscopy-positive subset | 46 | **Pv+** |  |  | + |  |  |  |
| Below 10 y.o. from Bataan | 202 | **neg** |  | — | — | — | — | — |
| Historical positives aged ≥50y.o. from Occidental Mindoro and Palawan | 711 |  |  | — |  | + |  |  |
| PfAMA1 and PfMSP1<sub>19</sub> seropositive subset | 512 |  |  |  |  |  | + |  |
| PvAMA1 and PvMSP1<sub>19</sub> seropositive subset | 324 |  |  |  |  |  |  | + |
| Randomly selected malaria-negatives from Palawan aged <50y.o.** | 550 |  |  | — | — |  |  |  |
| **Bataan study**<sup>30</sup> |  |  |  |  |  |  |  |  |
| Below 10 y.o. from Bataan | 73 | **neg** |  | — | — | — | — | — |
| **Malaysia study**<sup>29</sup> |  |  |  |  |  |  |  |  |
| *P. falciparum*-positive subset | 17 | **Pf+** |  | + |  |  |  |  |
| *P. vivax*-positive subset | 37 | **Pv+** |  |  | + |  |  |  |
| Historical positives aged ≥50y.o. from Malaysia 2 | 1581 |  |  |  |  | + |  |  |
| PfAMA1 and PfMSP1<sub>19</sub> seropositive subset | 1479 |  |  | — |  |  | + |  |
| PvAMA1 and PvMSP1<sub>19</sub> seropositive subset | 873 |  |  | — |  |  |  | + |

\* Finite mixture models (FMM) were derived from the entire ENSURE dataset (n=9132), and indicated validation dataset was used to determine receiver operating characteristics (ROC) curves for both FMM and Negative population models (seropositivity classifications from single antigens). For the supervised classification models, + means it was classified as positive for the model, and — means it is considered for negative classification. Abbreviations: Pf- *P. falciparum*, Pv- *P. vivax*, neg – malaria-negative; y.o.-years old

### Applying machine learning techniques for multiplex analysis of antigen responses

We next utilized machine learning (ML) techniques on multiplex serological data for determining recent and historical malaria exposure. The 8 Pf-specific antigens along with age were the covariates evaluated for *P. falciparum* exposure, while for *P. vivax* exposure, only the 6 Pv-specific antigens were analyzed (age data was not available). The training data used for these models are in Table 2. To account for the age-dependent cumulative increase in antibodies, the negative dataset for classifying recent infection included historical positives – survey participants aged >50 years from Malaysia^29^ and Philippines^30^ assumed to have had historical exposure, as well as a random selection of Pf or Pv-negatives from all age groups from this current study. For classifying historical exposure, the training dataset included the historical positives as positives, and the same negatives from the validation dataset (Table 2).

Super Learner (SL) optimized with AUC (Area under the ROC Curve) was used as an ensemble modelling algorithm to allow for evaluating multiple models simultaneously^23,31,32^, namely Random Forest (RF), k-Nearest Neighbor (kNN), generalized boosted models (GBM), Support Vector Machine (SVM), and GLM with Lasso (glmnet). Feature selection with corP, that screens for univariate correlation, was also included for some component models (GBM, RF). The SL model gives a prediction value that ranges from 0 to 1, and this can be used to obtain binary classification (*i*.*e*., those with prediction values higher than 0·5 were considered positive). A 20-fold cross-validation was performed for internal validation to evaluate the performance of the fitted SL models using a withheld training dataset, and the whole training dataset. ROC curves were used to evaluate the outcome of predicted values for the samples and were then compared to single antigen performances. The performance of each ML model, or base learner, in the SL ensemble was also assessed. For additional assessment, external validation was performed for the classification models through an independently collated dataset, which used the same panel of *P. falciparum* antigens (not available for *P. vivax* panel).

### Evaluating model performance in identifying current Pf and Pv infections in this study

Using this study’s malaria-positive samples, we assessed the capability of the serological markers in identifying falciparum and vivax infections (Supplementary Table 1). The single antigen performances were compared with SL predictions, where models included 3 to 9 covariates combined based on the variable importance outcomes. The AUC values from the ROC analysis based on training data and ENSURE malaria-positives (test data) were then computed. With Super Learner giving weights to multiple ML models simultaneously, we also evaluated the individual ML models that were given the most weight or importance, namely RF and GBM.

### Estimating seroconversion rates and historical exposure

Seropositivity of the cumulative exposure markers (AMA1, MSP1_19_ for both *P. falciparum* and *P. vivax*) were used to estimate seroconversion rates (SCR) and seroreversion rates (SRR), by fitting the age-specific prevalence for each study site into reverse catalytic models using likelihood ratio methods, as previously described^3,33^. The predicted time of change in transmission is analyzed. In addition to the AMA1 and MSP1_19_ seroprevalence based on FMM models, RF classifications using the combined AMA1 and MSP1_19_ data for both *P. falciparum* (Pf-RF:2-covar) and *P. vivax* (Pv-RF:2-covar) were also used to generate seroprevalence curves. The positive training data used for these RF models are subsets of the historical positives that were seropositive to both markers for each species based on FMM cutoff values (Table 2). The Super Learner approach was also used to predict Pf and Pv historical exposure using multiplex antibody data.

## Results

### Heterogeneity of antibody responses from the 3 collection sites

A total of 9132 DBS samples (6572 for Palawan, 1683 for Occidental Mindoro, and 877 for Bataan) were available for serological evaluation. Females comprised >60% of survey participants, while all age groups were well-represented (Table 3). Malaria infections were detected only in Palawan, with 51·8% and 48·2% of infected males and females, respectively, aged under 10 years old. Of the 889 *Plasmodium-*positive samples confirmed through either microscopy, RDT and/or PCR, 58·0% had *P. falciparum*, 12·4% had *P. vivax*, 6·7% had mixed Pf+Pv infections, 6·1% had *P. malariae, P. ovale* or *P. knowlesi*, while species identification could not be confirmed for 16·8% of the PCR-positive samples. From the Pf and Pv malaria-positive individuals (n=707, including mixed infections), 388 (54·8%) had fever before or during the health facility visit, 62·1% of which were ≤10 years old. Conversely, 63·1% of the asymptomatic malaria infections were >10 years old.

**Table 3.** Characteristics of study population by site

|  | Morong, Bataan | Abra de Ilog, Occidental Mindoro | Rizal, Palawan |
| --- | --- | --- | --- |
| Number of analyzed samples | **877** | **1683** | **6572** |
| Female, % | **628 (71.6%)** | **1101 (65.4%)** | **3734 (56.8%)** |
| Age, median (IQR) | **26 (11-39)** | **28 (15-43)** | **13 (5-31)** |
| number (% in site) |  |  |  |
| 0 to 5 | 150 (17.1%) | 150 (8.9%) | 1900 (28.9%) |
| 6 to 10 | 63 (7.2%) | 154 (9.2%) | 1055 (16.1%) |
| 11 to 19 | 102 (11.6%) | 234 (13.9%) | 957 (14.6%) |
| 20 to 34 | 279 (31.8%) | 508 (30.2%) | 1222 (18.6%) |
| 35 up | 283 (32.3%) | 637 (37.8%) | 1438 (21.9%) |
| *Plasmodium*-positive by microscopy/RDT/PCR (% in site)* | **0 (0.0%)** | **0 (0.0%)** | **889 (13.5%)** |
| *P. falciparum*-positive (by microscopy/RDT/PCR)* | **0 (0.0%)** | **0 (0.0%)** | **595 (9.1%)** |
| *P. vivax*-positive (by microscopy/RDT/PCR)* | **0 (0.0%)** | **0 (0.0%)** | **172 (2.6%)** |
| PCR-confirmed species ID, n (% in site) |  |  |  |
| *P. falciparum*, Pf mono-infection | 0 (0.0%) | 0 (0.0%) | 516 (7.9%) |
| *P. vivax*, Pv mono-infection | 0 (0.0%) | 0 (0.0%) | 110 (1.7%) |
| Pf + Pv mixed infection | 0 (0.0%) | 0 (0.0%) | 60 (0.9%) |
| Other species (mono- or mixed infections) | 0 (0.0%) | 0 (0.0%) | 55 (0.8%) |
| With fever or history of fever (% in site) | **69 (7.9%)** | **172 (10.2%)** | **2727 (41.5%)** |
| Symptomatic Pf and Pv infections* | 0 (0.0%) | 0 (0.0%) | 388 (5.9%) |
| Asymptomatic infections (no fever, *Plasmodium*-positive)* | **0 (0.0%)** | **0 (0.0%)** | **428 (6.5%)** |
| Asymptomatic microscopy-positive Pf infections | 0 (0.0%) | 0 (0.0%) | 132 (2.0%) |
| Asymptomatic microscopy-positive Pv infections | 0 (0.0%) | 0 (0.0%) | 35 (0.5%) |

\* Numbers include all *Plasmodium* infections detected by microscopy, RDT and/or PCR. Some samples did not have enough material for further species identification by PCR, for which results were based on microscopy and/or RDT diagnosis, while some (n=149) were confirmed *Plasmodium*-positive only. Pf and Pv numbers reported include mixed infections, which were specified in the PCR-confirmed species ID breakdown.
\*\* All 5 *Plasmodium* species, including *P. malariae* (n=50), *P. ovale* (n=4) and *P. knowlesi* (n=1), were detected in Palawan, with some as co-infections with Pf and/or Pv.

Firstly, we compared the magnitude of antibody responses to *P. falciparum* and *P. vivax* antigens from the 3 collection sites. Palawan consistently had the highest antibody levels to all antigens and in all age groups, followed by individuals from Occidental Mindoro then Bataan (Fig 2A, Supplementary Figure 1). Expectedly, cumulative exposure markers PfAMA1, PfMSP1_19_, PfGLURP R2, PvAMA1 and PvMSP1_19_ were strongly correlated with age (Spearman’s coefficient >0·5, *p*<0·0001). Recent exposure markers Etramp5.Ag1 and GEXP18 were less strongly associated (Spearman’s coefficient of 0·39 and 0·42, respectively, p<0·0001). PvMSP1_19_ and PvDBPRII showed weak correlation (Spearman’s coefficient of 0·13 and 0·12, respectively, p<0·0001) with current *P. vivax* infection (Figure 2B). Similarly, mean antibody levels were higher in a species-specific manner (Figure 3). Taken together, our results confirm the applicability of these serological markers in differentiating areas of varying malaria endemicity, and there were species-specific associations with current infection.

**Figure 2.**
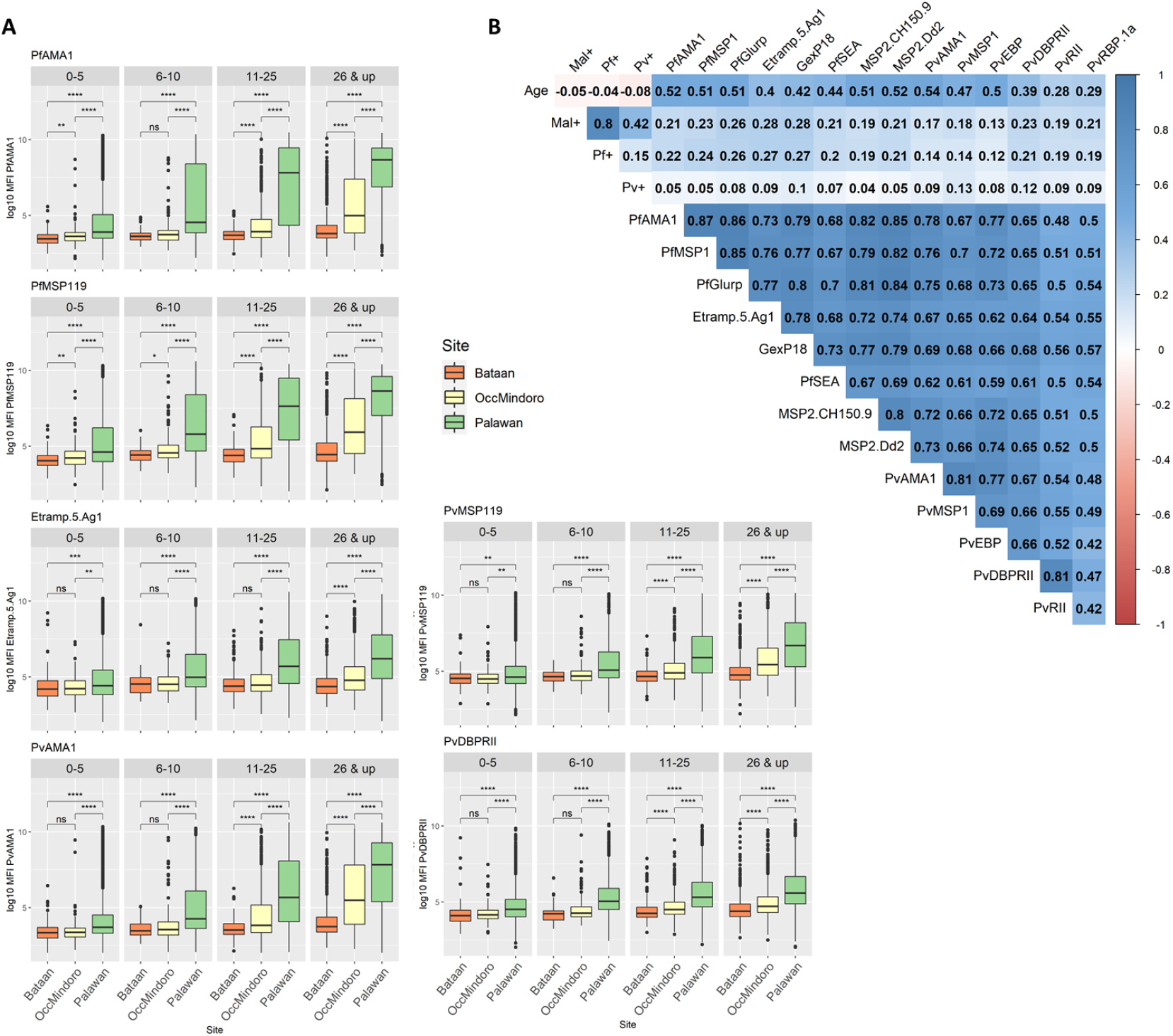
Antibody responses to serological markers of *P. falciparum* and *P. vivax* correlate with malaria incidence. **A.** Antibody levels (reported as log10 MFI) in response to *P. falciparum* cumulative and recent exposure markers PfAMA1, PfMSP1_19_, and Etramp5.Ag1, and *P. vivax* serological markers PvAMA1, PvMSP1_19_ and PvDBP.RII by study site and age group. Statistical difference of overall antibody responses among study sites within age groups were determined using Kruskall-Wallis test and Wilcoxon test for pairwise comparisons (*p < 0·05, **p < 0·01, ***p < 0·001, ****p < 0·0001). **B.** Spearman’s correlation coefficients for age, malaria diagnosis (Mal+: Plasmodium-positive, Pf+: *P. falciparum*-positive, Pv+: *P. vivax*-positive), and antibody responses to the 14 antigens in the panel for Palawan samples (n=6572).

**Figure 3.**
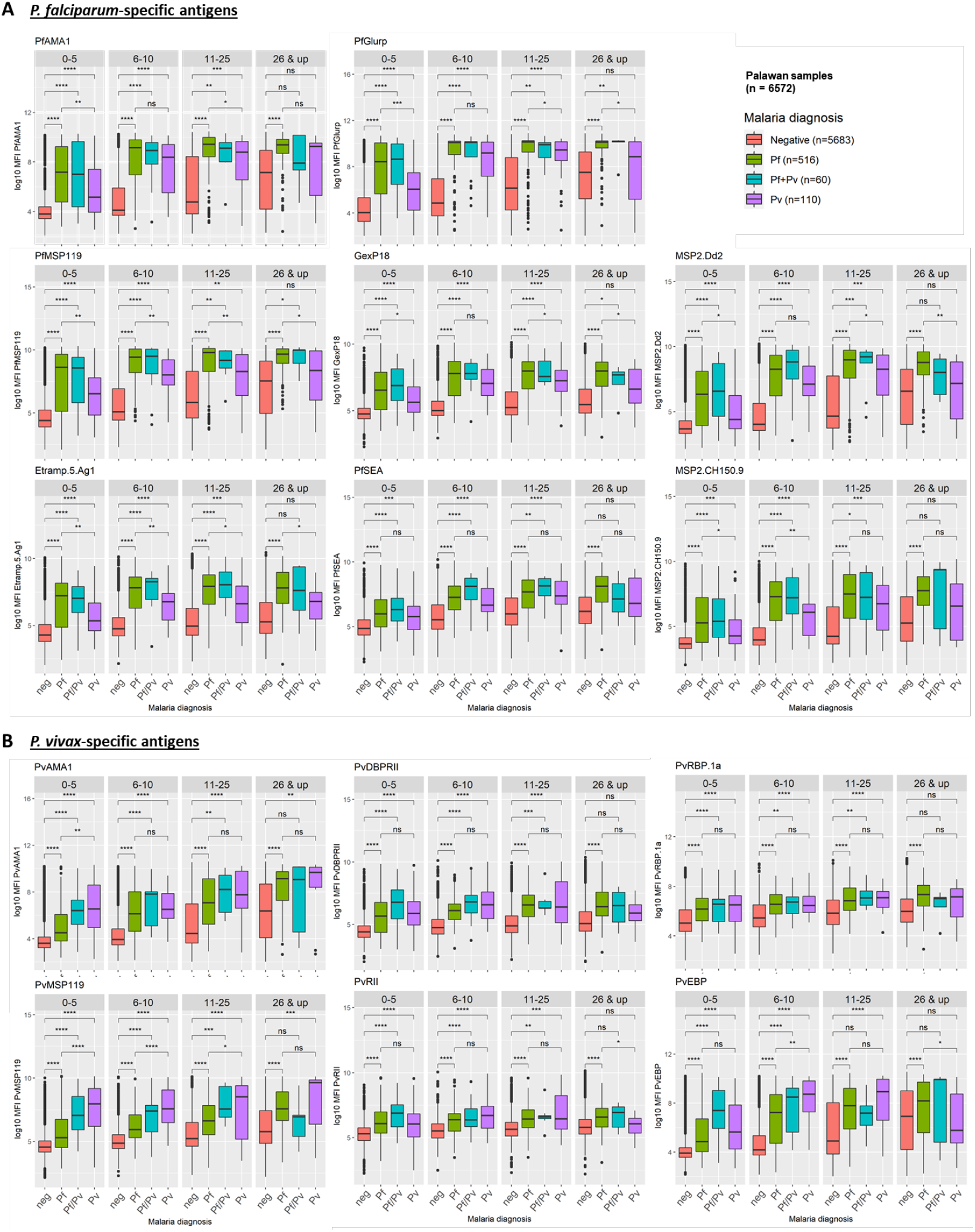
Serological markers exhibit species-specific association with current infection in varying levels. **A-B)** Comparison of antibody titers of Palawan samples (n=6572) for *Plasmodium*-positive (Pf: P. falciparum-positive, Pf/Pv: Pf and Pv mixed infection, Pv: P. vivax-positive) and negative (neg) samples by age group in each *P. falciparum*-specific (A) and *P*.*vivax*-specific antigen.

### Identifying markers of recent falciparum and vivax malaria exposure and current infection

Next, we applied different analytical approaches to ascertain whether the serological markers can be used to predict recent or historical exposure. The resulting seropositivity cutoff values were comparable for FMM (unsupervised) and Negpop (supervised) classification models, with AUC values from the validation data ranging from 0·812 to 0·932 for the 8 *P. falciparum* markers, and 0·534 to 0·943 for the 6 *P. vivax* markers (Figure 4A & 4B, Supplementary Tables 1 & 2). When applied to classifying study samples, some of the markers showed low predictive ability, with AUC values ranging from 0·497 to 0·756 for *P. falciparum* markers, and 0·513 to 0·731 for *P. vivax* markers. Etramp5.Ag1 and GEXP18 for *P. falciparum*, and PvMSP1_19_ for *P. vivax*, had significantly higher AUC values (>0·735, *p*<0·002) compared to the rest of the markers analyzed (Supplementary Tables 1 & 2). Density plots of antibody responses also show overlap of malaria-negative and positive distributions, contributing to the lower AUCs (Supplementary Figure 2). Seroprevalence rates for all antigens were consistently low in Bataan in all age groups (Figures 4A & 4B), with recent exposure markers (hollow circles) estimating lower rates.

**Figure 4.**
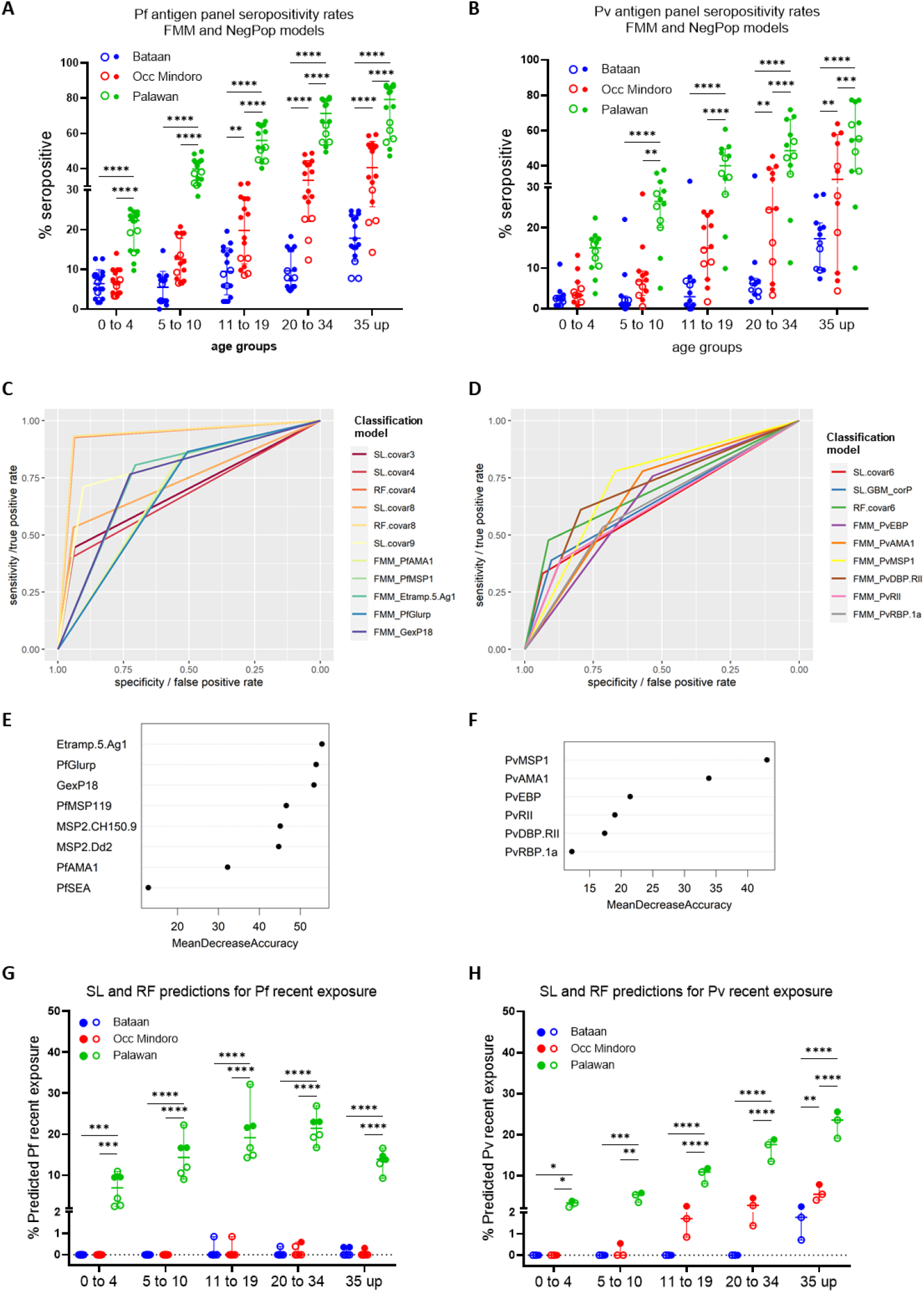
Analysis of serological markers through machine learning methods improves classifications for recent *P. falciparum* and *P. vivax* exposure or current infection. **A-B**. Seropositivity rates of sample population by site and age group based on cutoff values from finite mixture models (FMM) and negative population model (NegPop) for Pf and Pv antigens (detailed in **Supplementary Tables 1 and 2**). Hollow dots represent the FMM and Negpop seropositivity rates for the two reported recent exposure markers Etramp5.Ag1 and GEXP18 for Pf (A), and PvMSP1_19_ and PvDBP.RII for Pv (B) panel. Lines with error bars represent median with 95% CI. **C-D**. Receiver operating characteristic (ROC) curves for the antibody responses to single antigens for individual antigens, and for the SL models (shown in both as binary outcomes of seropositivity / prediction values). **E-F**. Variable importance of the 8 Pf-specific (E) and 6 Pv-specific (F) antigens based on the Random Forest model. **G**. Predicted rates of recent Pf exposure based on analysis of the continuous antibody responses of the 8-antigen panel using Super Learner. Each hollow dot represents differences in the number of covariates used for the model (3, 4, 8, 9), as well as 2 showing rates from prediction values of the Random Forests (RF) component model (RF.covar4 and RF.covar8 in solid dots). **H**. Predicted rates of recent Pv exposure based on analysis of the continuous antibody responses of the 6-antigen panel using Super Learner. Dots represent the positivity rates from the prediction values of the SL model, and the individual predictions from the 2 most weighted base learners in the resulting model – RF and GBM. RF predictions (RF.covar6) are shown as solid dots. (SL: Super Learner, RF: random Forest, covar#: number of covariates included in the model, FMM: finite mixture models; *p < 0·05, **p < 0·01, ***p < 0·001, ****p < 0·0001 with significance assessed by one-way ANOVA followed by Tukey’s multiple comparison)

We next applied ML methods for analyzing multiplex antibody response data. The Super Learner (SL) ensemble machine learning algorithm was used to simultaneously evaluate classifications from ML models, which included Random Forest (RF), k-Nearest Neighbor (kNN), generalized boosted models (GBM), Support Vector Machine (SVM), and GLM with Lasso (glmnet), and weights are applied to each learner after cross-validation. Covariates were assessed for their relative importance for the model, and different combinations in a model were assessed using the ROC curves of cross-validated predictions (Figure 4C & 4D; Supplementary Tables 1 & 2). The 9-covariate model (SL: 9-covar; 8 antigens and age as covariates) had the highest AUC among all SL models, with a value of 0·9898 (95% CI 0·9874-0·9921) and 0·9197 (95% CI 0·9093-0·9302) for the binary classification predictions for the training data and test data, respectively, and removing the age as covariate (SL:8-covar model) decreased the prediction capacity (Supplementary Table 1). For predicting *P. vivax* recent exposure, the model incorporating the 6 Pv antigens as covariates (SL:6-covar) had a resulting AUC of 0·8857 (95% CI 0·8429-0·9284) and 0·6332 (0·5979-0·6686) for validation data and test data, respectively (Supplementary Table 2).

After a 20-fold nested cross-validation, generalized boosted models with or without feature selection (GBM and GBM_corP) were given the highest weights in the SL ensemble models for both species’ recent exposure predictions (Supplementary Figure 3A & 3B). However, upon closer inspection of the classification accuracy of the individual learners for predicting *P. falciparum* recent infections, the Random Forest (RF) models were found to have the highest AUCs in both the validation data (not shown) and test data (Supplementary Figure 3C). Computing for the AUCs of resulting binary classifications (Figure 4C) also showed RF models with highly improved AUCs compared to FMM-based single antigen classifications and even the SL models, suggesting that RF is the best performing model in distinguishing current or recent falciparum malaria infections. On the other hand, vivax infection classification models had comparable AUCs ranging from 0·701 to 0·8 for test data (Supplementary Figure 3D), and also for binary classifications for SL, RF and GBM models (Figure 4D; Supplementary Table 2).

Variable importance in the RF model for *P. falciparum* positivity (Figure 4E) shows that responses to Etramp5.Ag1, PfGLURP R2, GEXP18 and PfMSP1_19_ had the highest influence in the predictions, while PvMSP1_19_ and PvAMA1 were the most predictive *P. vivax* antigens (Figure 4F). RF.covar4 gave an AUC of 0·9983 for the training data and 0·9591 for the test data (Figure 4C; Supplementary Table 1), and had comparable AUC with the RF 8-covariate model (RF.covar8), showing that analysis using the RF model combining just the 4 predictive Pf antigens was sufficient for predicting recent *P. falciparum* infection or exposure.

### Model validation using external datasets

To further evaluate the performance of the RF:4-covar model for predicting *P. falciparum* recent exposure, external validation was performed using a collated test dataset (Supplementary Table 3) that included samples from a cross-sectional survey in Malaysia^34^ (n=8163; all Pf-negative), a 2017 cross-sectional survey in Bataan^30^ (n=1926; all malaria-negative), PCR-validated malaria cases (Pf=47, non-Pf=428), and malaria-naïve or malaria-negative samples (n=506). The model prediction resulted in an AUC of 0·93 (CI 0·8817-0·9782), and was able to predict 0·47% prevalence of infection for the Malaysia cross-sectional survey, and 0·10% for the Bataan cross-sectional survey. It also correctly identified 41 of 47 (87·2%) of the *P. falciparum* PCR-confirmed cases from the malaria-positive subset.

### Estimating P. falciparum and P. vivax historical exposure

AMA1 and MSP1_19_ responses for *P. falciparum* and *P. vivax* have been widely used to assess historical exposure by estimating seroconversion and seroreversion rates. Using reverse catalytic models and maximum likelihood tests on the age-specific seroprevalence based on the FMM model seropositivity rates for PfAMA1, PfMSP1_19_, PvAMA1 and PvMSP1_19_, we estimated the time of interruption of transmission in our study area (Figure 5, Table 4). The RF algorithm was also employed to generate models that combine both antigens (2-covariate models Pf-RF:2-covar and Pv-RF:2-covar), and the predicted binary outcomes were also used in the reverse catalytic models. Most of the seroconversion curves were best fit with a model assuming a change in transmission based on log likelihood tests (*p* < 0·001), except for PvMSP1_19_ (p= 0·507) and the Pv-RF:2-covar (p=0·285) model (Table 4). Based on these models, the time of change in transmission in Bataan was estimated at 29-33 years for *P. falciparum* and 28-29 years for *P. vivax*. Occidental Mindoro had varied estimates of the change point (7 and 12 years). For these 2 provinces, the seroconversion rates suggest a decrease in transmission, while an increase of the seroconversion rates for Palawan after the 2-year change point, suggested an increase in transmission in this study site.

**Table 4.** Seroconversion and seroreversion rates from reverse catalytic models in each study site

|  |  | Bataan | Occidental Mindoro | Palawan |
| --- | --- | --- | --- | --- |
| PfAMA1 | SCR<sub>historical</sub> | 0.02102 (0.00599 - 0.07375) | 0.03306 (0.02150 - 0.05082) | 0.06953 (0.06251 - 0.07736) |
|  | SCR<sub>recent</sub> | 0.00193 (0.00112 - 0.00332) | 0.01024 (0.00707 - 0.01483) | 0.13346 (0.11802 - 0.15092) |
|  | SRR | 0.010002 (0.0010 - 0.10353) | 0.01239 (0.00575 - 0.02673) | 0.00841 (0.00626 - 0.01129) |
|  | Time of change in transmission (yrs) | 29 | 12 | 2 |
| PfMSP1<sub>19</sub> | SCR<sub>historical</sub> | 0.04042 (0.00836 - 0.19516) | 0.03399 (0.02221 - 0.05204) | 0.07521 (0.06658 - 0.08495) |
|  | SCR<sub>recent</sub> | 0.00243 (0.00148 - 0.00396) | 0.01412 (0.00842 - 0.02367) | 0.14811 (0.13109 - 0.16734) |
|  | SRR | 0.01959 (0.00564 - 0.06796) | 0.01779 (0.00926 - 0.03416) | 0.01221 (0.00946 - 0.01575) |
|  | Time of change in transmission (yrs) | 33 | 7 | 2 |
| RF with PfAMA1 and PfMSP1<sub>19</sub> | SCR<sub>historical</sub> | 0.01513 (0.00426 - 0.05370) | 0.03338 (0.02172 - 0.05130) | 0.06836 (0.06133 - 0.07619) |
|  | SCR<sub>recent</sub> | 0.00122 (0.00064 - 0.00234) | 0.00931 (0.00630 - 0.01377) | 0.12847 (0.11333 - 0.14564) |
|  | SRR | 0.00463 (0.00002 - 0.91264) | 0.01327 (0.00639 - 0.02755) | 0.00901 (0.00674 - 0.01204) |
|  | Time of change in transmission (yrs) | 29 | 12 | 2 |
| PvAMA1 | SCR<sub>historical</sub> | 0.07078 (0.01619 - 0.30943) | 0.02760 (0.02094 - 0.03638) | 0.05210 (0.04652 - 0.05834) |
|  | SCR<sub>recent</sub> | 0.00197 (0.00102 - 0.00377) | 0.00840 (0.00451 - 0.01564) | 0.08800 (0.07591 - 0.10202) |
|  | SRR | 0.03138 (0.01524 - 0.06460) | 0.00715 (0.00269 - 0.01898) | 0.01074 (0.00801 - 0.01441) |
|  | Time of change in transmission (yrs) | 29 | 7 | 2 |
| PvMSP1<sub>19</sub> | SCR<sub>historical</sub> | 0.02858 (0.00465 - 0.17570) | 0.01143 (0.00921 - 0.01419) | 0.03760 (0.03172 - 0.04458) |
|  | SCR<sub>recent</sub> | 0.00178 (0.00092 - 0.00347) |  | 0.08687 (0.07507 - 0.10052) |
|  | SRR | 0.02941 (0.00862 - 0.10026) | 0.00404 (0.00047 - 0.03486) | 0.02146 (0.01599 - 0.02880) |
|  | Time of change in transmission (yrs) | 28 | - | 2 |
| RF with PvAMA1 and PvMSP1<sub>19</sub> | SCR<sub>historical</sub> | 0.03387 (0.00559 - 0.20521) | 0.01139 (0.00919 - 0.01414) | 0.04023 (0.03472 - 0.04661) |
|  | SCR<sub>recent</sub> | 0.00169 (0.00084 - 0.00340) |  | 0.08479 (0.07313 - 0.09833) |
|  | SRR | 0.03498 (0.01265 - 0.09673) | 0.00388 (0.00042 - 0.03598) | 0.02227 (0.01674 - 0.02963) |
|  | Time of change in transmission (yrs) | 28 | - | 2 |

Seroconversion rates (SCR) and seroreversion rates (SRR) are presented for *P. falciparum* and *P. vivax* based on reverse catalytic models using age-specific seroprevalence from finite mixture models with AMA1 and MSP1<sub>19</sub> and Random Forest (RF) 2-covariate models. If the best reverse catalytic model to fit the age-adjusted seropositivity data was one with no change point in transmission, only 1 SCR is indicated, while if the best fit is one with a change point, the SCR before the change point is indicated as SCR<sub>historical</sub> and the SCR after the change point is indicated as SCR<sub>recent</sub>. The estimated change point in transmission (in years) is also indicated.

**Figure 5.**
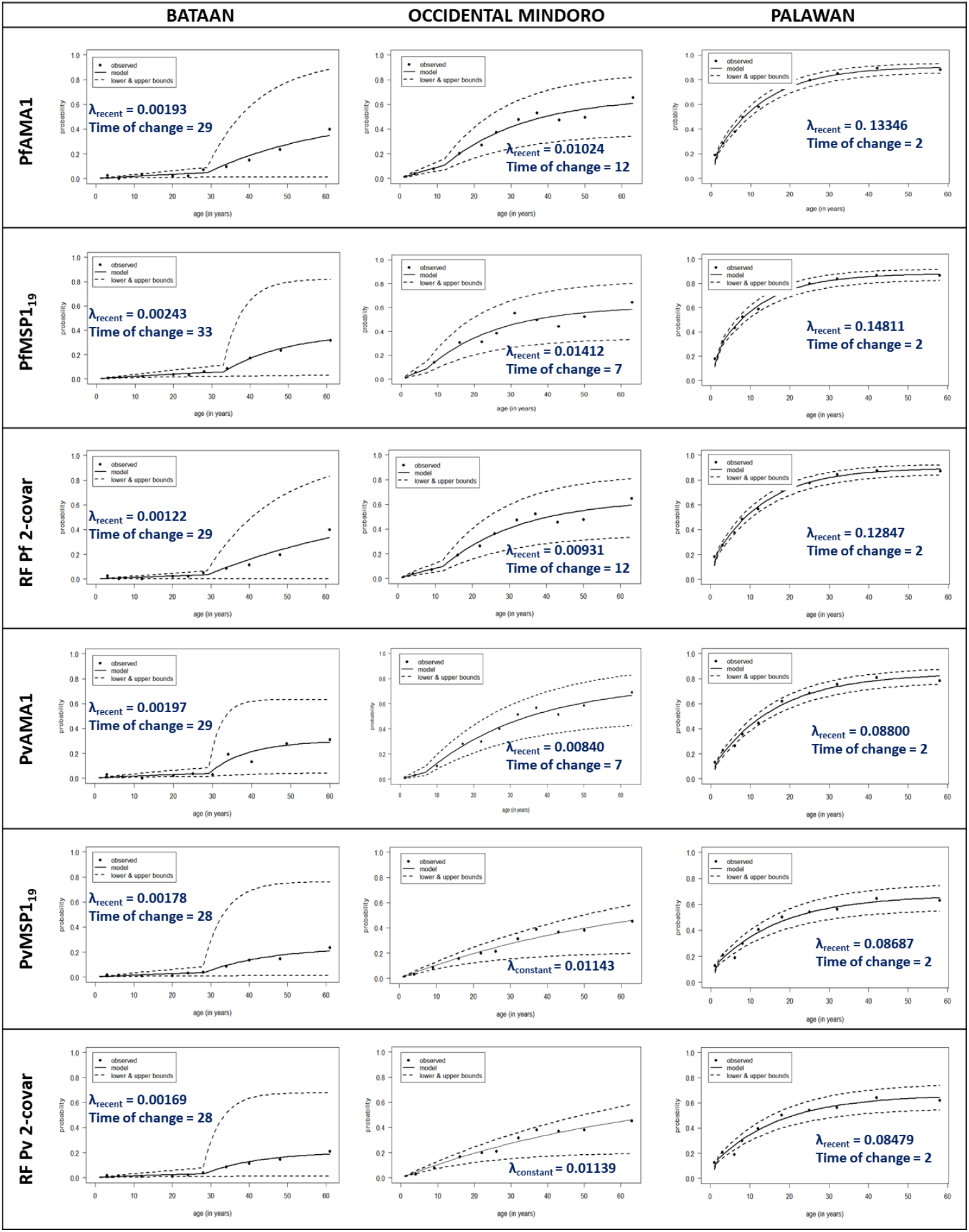
Seroconversion curves based on reverse catalytic models using AMA1 and MSP1_19_ antibody responses provide accurate estimates of historical exposure. Age-specific seroprevalence was based on finite mixture models and Random Forest models (using both antigens: RF 2-covar models) for each species. Solid lines represent the fit of the reversible catalytic models, dashed lines represent 95% confidence intervals, and dots represent the observed proportions of seropositives per age divided into 10% centiles. For models assuming a change point in transmission, only the recent seroconversion rates and change point estimates (in years) are shown, while the historical seroconversion rates and seroreversion rates are detailed in Table 4.

We further explored whether the use of SL model can provide improved estimates of historical exposure. The exposure prediction values from the SL model (8 *P. falciparum* markers) were graphed for each sample by site and age (Figure 6A & 6D), and compared with seropositivity rates of PfAMA1 (Figure 6C), which is shown to have the highest influence in the RF model prediction (Figure 6B). However, comparing the SL-predicted rates with AMA1 and MSP1_19_ seropositivity rates (Figure 6D vs 6E) show that SL overestimated historical exposure, with higher-than-expected positivity rates in younger age groups from Bataan and Occidental Mindoro, where transmission is reportedly absent. The seropositivity rates based on the 2-covariate RF models with AMA1 and MSP1_19_ seem to provide better estimates of historical exposure to both *P. falciparum* and *P. vivax* based on known malaria prevalence of the sites than the SL models (Figure 6E & 6F).

**Figure 6.**
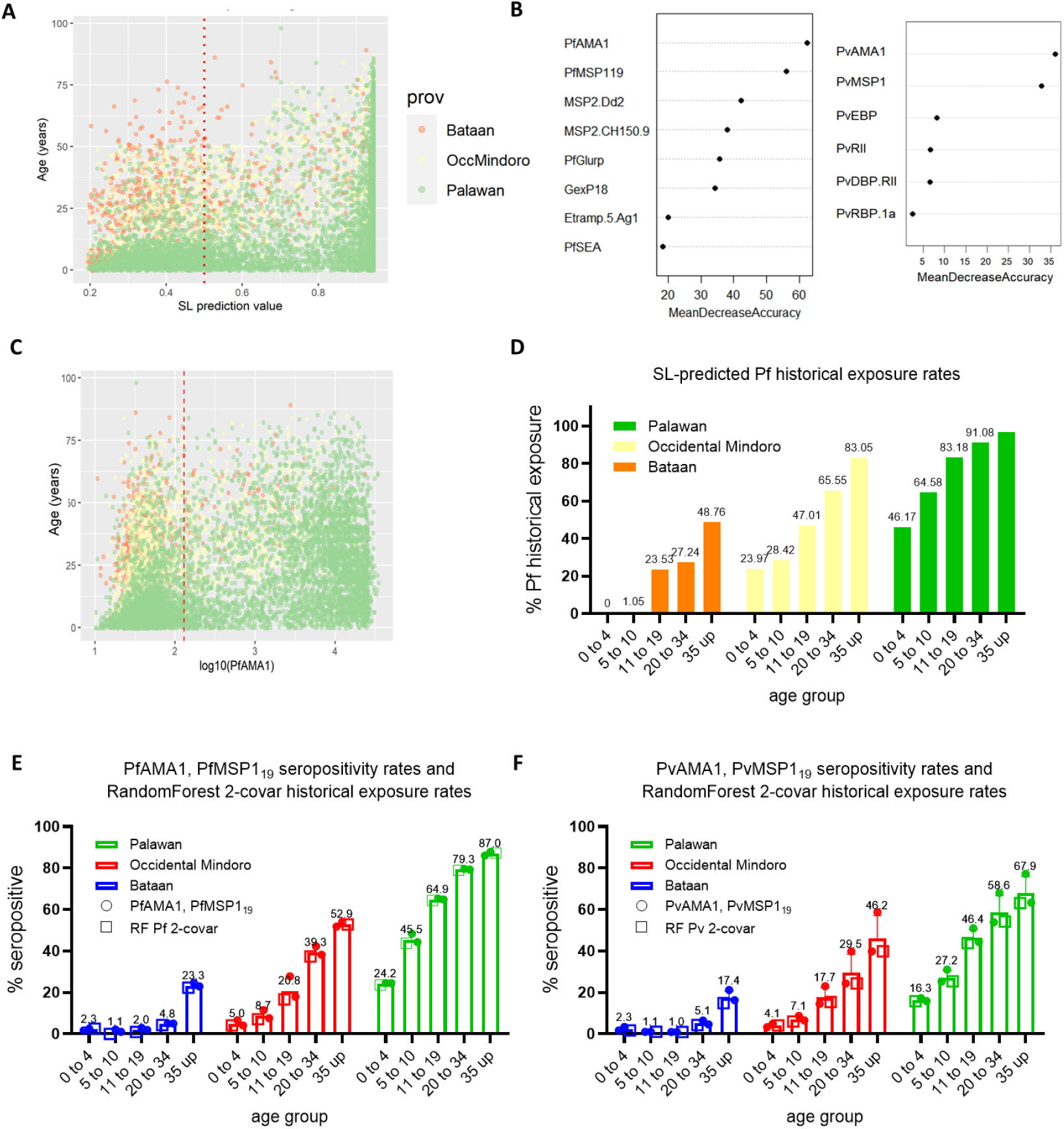
Cumulative exposure markers confirm historical *P. falciparum* and *P. vivax* exposure, and heterogeneity of transmission in the 3 sites. **A**. Plot of Super Learner prediction values for Pf historical exposure by site and age, using the model with 8 Pf-specific serological markers as covariates. Red dotted line represents positivity cutoff at 0·5. **B**. Variable importance based on the Random Forest model of the 8 Pf-specific antigens and 6 Pv-specific antigens in predicting historical exposure for each species. **C**. Distribution of antibody responses to PfAMA1 by site and age of individuals (n=9132). Red dashed line represents the seropositivity cutoff value from the FMM model. **D-E**. Summary graphs per age category per site of SL-predicted Pf historical exposure (D) and PfAMA1, PfMSP1 seropositivity rates graphed with estimated historical exposure rates using the Random Forest model with PfAMA1, PfMSP119 as covariates (E). **F**. Summary graph of PvAMA1, PvMSP1 seropositivity rates with estimated historical exposure rates using the Random Forest model with PvAMA1, PvMSP119 as covariates seropositivity per age category per site

## Discussion

In this study we assessed the utility of multiple antigen-specific antibody responses and different statistical methods to estimate both current malaria infection and historical transmission in areas of different endemicity in the Philippines. Using a multiplex bead-based assay with a panel of 8 *P. falciparum*-specific and 6 *P. vivax*-specific recombinant antigens on DBS samples, we showed how the use of combined antibody data can improve predictions of recent or historical malaria exposure. In particular, machine learning model predictions of malaria exposure were consistent with reported levels of transmission in our sites, which highlight the potential for advanced analysis of multiplex serological data to provide accurate complementary data on incidence levels that could be used by control programs at small spatial scales.

In many countries including the Philippines, malaria-endemic provinces apply for malaria-free status when indigenous cases have not been reported for a set number of years, typically 3 or more. More rapid, subnational demonstration of the absence of exposure could aid in this process. The use of multiple antigens to assess exposure to malaria infection circumvents some issues related to genetic diversity in the parasite and variation in the human immune response to different antigenic targets. The approach also allows application of more advanced statistical analysis to examine optimal combinations of antibody responses in predicting specific outcomes. Here, in addition to the well-studied antigens AMA1 and MSP1_19_, we screened other antigenic markers that have been reported to accurately predict recent exposure in studies in Africa and the Caribbean^14,23,36^. PfGLURP R2, Etramp5.Ag1 and GEXP18 have been identified as recent exposure markers for *P. falciparum*^23,36^ and PvMSP1_19_ for *P. vivax*^25^. Our RF variable importance results also identified these markers to be the most predictive in our panel.

In assessing the individual performances of the serological markers, we show that the antibody responses to all antigens were species-specific, consistent with previous observations^37,38^. For some recent exposure makers, we observed <10% seropositivity in younger age groups from malaria-free Bataan, suggesting background seropositivity. When we employed machine learning^23,25,39^, our models were able to confirm the absence of current infection in Occidental Mindoro and Bataan (although there was lower precision for *P. vivax* exposure prediction), and, using the Random Forest predictions from a 4-covariate model including PfGLURP R2, Etramp5.Ag1, GEXP18 and PfMSP1_19_, accurately identified >92·0% of the *Plasmodium-*positive study samples from this study, whether detected through microscopy, RDT and/or PCR. It also correctly identified 87·2% of *P. falciparum* positives from a Malaysian dataset. The observation that this relatively simple model, which did not require epidemiological variables such as age, was able to provide accurate predictions for different datasets, shows its potential as a robust indicator for recent exposure and current infections, in areas with varying levels of transmission.

We expected that the Super Learner model would improve the predictions with its ensemble approach; however, for predicting *P. falciparum* current infections in particular, RF-based models had the better AUC for both training and test data. This was not the case for the *P. vivax* predictions, which showed similar AUCs (<0·7) for the final SL model and its component models. It is likely that the low predictive ability we observed from the *P. vivax* panel is a limitation of the panel itself rather than the analytical approaches used. Notwithstanding, the SL approach was able to provide the means for evaluating multiple supervised ML models simultaneously, and, for the purpose of accurately distinguishing recent exposure from historical exposure with high sensitivity, the RF model seems to provide accurate estimates.

This study had other limitations. For classifying recent and historical exposure, we had no data on the malaria history of the survey participants, such that we instead focused on predicting for current infections. Still, external validation of our prediction algorithm for *P. falciparum* infection confirms its promising performance in accurately predicting malaria infections. Also, since the study analyzed samples from a health facility-based survey, whether or not it accurately represents the populations needs to be assessed. The bias towards sampling health-seeking individuals was partially addressed by also recruiting companions of patients who visited the health facilities ^26^. When we used our RF.covar4 model to predict recent exposure in the 2017 cross-sectional survey also conducted in Morong, Bataan, it also predicted 0·10% recent exposure – similar with our health facility-based data. The 29 to 30-year change point in transmission for Bataan can also be deemed consistent with the reported change point at 22 years in this previous survey^30^.

Although we cannot validate the predicted historical exposure, we are able to show that consistent with previous studies^10,21,24,40,41^, AMA1 and MSP1_19_ seropositivity rates (individually or combined) for *P. falciparum* and *P. vivax* seemed to agree with the known malaria situation of our study sites. The seroconversion rates also reflect this, with Bataan having the lowest, followed by Occidental Mindoro, and highest for Palawan. Our results are able to confirm the absence of *P. falciparum* and *P. vivax* transmission in malaria-free Bataan for the age groups 10 and below, providing evidence for its malaria elimination status. For Occidental Mindoro, the interruption in transmission from 7-12 years ago may be explained by the extensive malaria control activities implemented through the Global Fund for Malaria since the 2000s. Our seroconversion curve results also suggest an increase in malaria transmission in Rizal, Palawan during the time of our survey (2016 to 2018), and upon checking the records of reported malaria cases, there was indeed a steady rise of the API in this municipality from 25·43 in 2014, 33·97 in 2015, to 46·09 in 2016 and 56·78 in 2018.

Despite the mentioned limitations of the study, our results clearly show the potential use of multiplex antibody responses and applications of machine learning approaches in assessing malaria transmission for countries aiming for malaria elimination. Of interest for countries aiming for elimination is evaluating the impact of a decrease in transmission on the immunity or vulnerability of a population, especially in differing endemic settings^42,43^. Immunological studies are needed to investigate this further. In a sub-national elimination setting such as the Philippines, both recent and historical *P. falciparum* and *P. vivax* exposure metrics were indicative of the absence of recent transmission in Bataan and Occidental Mindoro, and also identified current infections in Rizal, Palawan, thus showing its ability in assessing the malaria situation in varying areas of endemicity. Our study provides baseline immunological data for monitoring risk populations in the Philippines. This serological surveillance approach can aid in devising control measures by malaria elimination programs, as well as provide evidence of the effectiveness of programs being implemented.

## Data Availability

All data analyzed in this study are available from authors upon reasonable request and with permission of relevant institutional review boards.

## Acknowledgements

This study was supported by the Newton Fund, Philippine Council for Health Research and Development, and UK Medical Research Council through funding received for the ENSURE project (MR/N019199/1). We also express our gratitude to the ENSURE team, especially Carol Joy Sarsadiaz, Hennessey Sabanal, Beaulah Boncayao and Ellaine de la Fuente, who assisted in the conduct of serological assays, as well as Dr. Mario Jiz, Dr. Catalino Demetria, and Dr. Marilou Nicolas who provided technical assistance and access to their Luminex 200 instruments. Also, we thank the local governments and health staff of Palawan, Occidental Mindoro and Bataan for their invaluable support to our research. This work is also supported by the Nagasaki University’s “Doctoral Program for World-leading Innovative and Smart Education” for Global Health, KENKYU SHIDO KEIHI.

## Author Contributions

CJD, FEJE, JCRH, and MLMM conceived and designed this study. MLMM, KMF and CJD analyzed and interpreted the data. MLMM wrote the first draft of the manuscript with support from KMF, CJD, JCRH, KY, and FEJE. JSL, RAR, MLMM, and APNB supervised the data and sample collection, and the conduct of assays. KKAT, JHA, CH, and CEC provided the recombinant antigens used for the multiplex bead-based assay. MLMM and RAR performed the serological assays with support from TH and KKAT. All authors read and approved the final manuscript.

---

**Supplementary Table 1.**
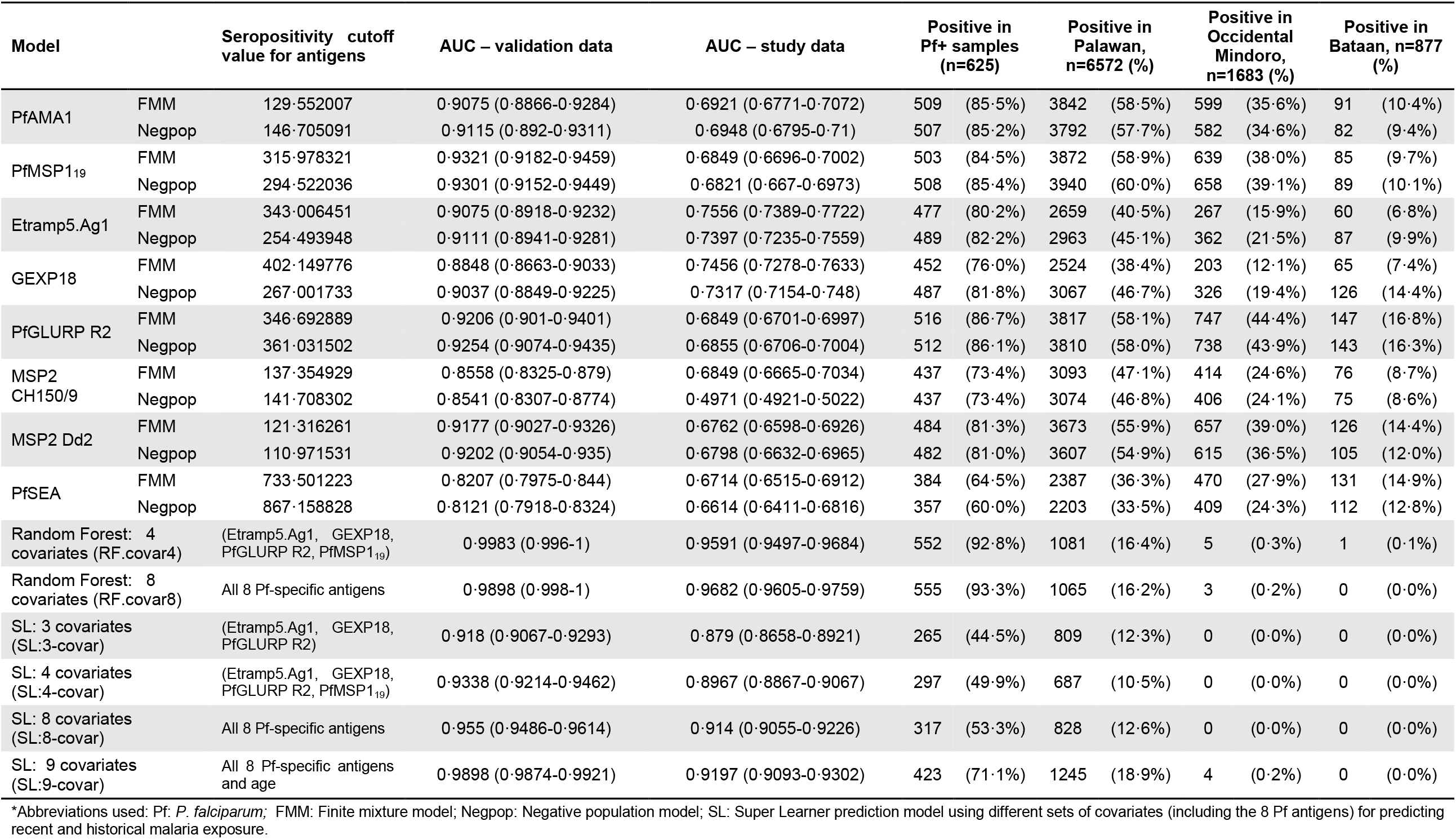
Seropositivity rates based on Pf-specific antigens and SuperLearner predictions for recent Pf exposure

**Supplementary Table 2.** Seropositivity rates based on 6 Pv-specific antigens and SuperLearner predictions for recent Pv exposure

| Model |  | Seropositivity cutoff value for antigens | AUC – validation data | AUC – study data | Positive in Pv+ samples (n=172) |  | Positive in Palawan, n=6572 (%) |  | Positive in Occidental Mindoro, n=1683 (%) |  | Positive in Bataan, n=877 (%) |  |
| --- | --- | --- | --- | --- | --- | --- | --- | --- | --- | --- | --- | --- |
| PvAMA1 | FMM | 149.776739 | 0.9185 (0.8934 - 0.9436) | 0.6744 (0.6424 - 0.7064) | 130 | (75.6%) | 2991 | (45.5%) | 650 | (38.6%) | 84 | (9.6%) |
|  | Negpop | 172.486298 | 0.9164 (0.8895 - 0.9434) | 0.6720 (0.6391 - 0.7049) | 127 | (73.8%) | 3035 | (46.2%) | 626 | (37.2%) | 77 | (8.8%) |
| PvMSP1<sub>19</sub> | FMM | 395.678911 | 0.9429 (0.9229 - 0.9629) | 0.7308 (0.6997 - 0.7618) | 134 | (77.9%) | 2588 | (39.4%) | 430 | (25.5%) | 63 | (7.2%) |
|  | Negpop | 666.248684 | 0.9234 (0.8935 - 0.9533) | 0.7049 (0.6692 - 0.7406) | 114 | (66.3%) | 2135 | (32.5%) | 295 | (17.5%) | 38 | (4.3%) |
| PvDBP.RII | FMM | 403.658933 | 0.8036 (0.7546 - 0.8527) | 0.7065 (0.6697 - 0.7434) | 105 | (61.0%) | 1699 | (25.9%) | 165 | (9.8%) | 50 | (5.7%) |
|  | Negpop | 285.09444 | 0.8143 (0.7668 - 0.8618) | 0.7037 (0.6681 - 0.7392) | 115 | (66.9%) | 2087 | (31.8%) | 221 | (13.1%) | 72 | (8.2%) |
| PvRBP.1a | FMM | 777.354467 | 0.7367 (0.6865 - 0.7869) | 0.6248 (0.5871 - 0.6625) | 92 | (53.5%) | 1979 | (30.1%) | 416 | (24.7%) | 242 | (27.6%) |
|  | Negpop | 2986.35245 | 0.5343 (0.5006 - 0.5679) | 0.5127 (0.4901 - 0.5352) | 17 | (9.9%) | 507 | (7.7%) | 104 | (6.2%) | 61 | (7.0%) |
| PvRII | FMM | 843.409764 | 0.7706 (0.7195 - 0.8218) | 0.6302 (0.5935 - 0.6670) | 67 | (39.0%) | 1079 | (16.4%) | 104 | (6.2%) | 38 | (4.3%) |
|  | Negpop | 377.466964 | 0.8007 (0.7530 - 0.8484) | 0.6322 (0.5955 - 0.6688) | 107 | (62.2%) | 2717 | (41.3%) | 489 | (29.1%) | 102 | (11.6%) |
| PvEBP | FMM | 147.668259 | 0.7735 (0.7247 - 0.8224) | 0.6471 (0.6144 - 0.6799) | 128 | (74.4%) | 3328 | (50.6%) | 719 | (42.7%) | 106 | (12.1%) |
|  | Negpop | 661.785062 | 0.7133 (0.6616 - 0.7649) | 0.6184 (0.5809 - 0.6559) | 101 | (58.7%) | 2608 | (39.7%) | 566 | (33.6%) | 63 | (7.2%) |
| SL: 6 Pv antigens |  | All Pv-specific antigens as covariates | 0.8857 (0.8429-0.9284) | 0.6332 (0.5979-0.6686) | 57 | (33.1%) | 603 | (9.2%) | 35 | (2.1%) | 2 | (0.2%) |
| RF: 6 Pv antigens |  |  | 1 | 0.6918 (0.6542-0.7293) | 81 | (47.1%) | 802 | (12.2%) | 57 | (3.4%) | 5 | (0.6%) |
| GBM: 6 Pv antigens |  |  | 0.9043 (0.8647-0.9439) | 0.6462 (0.6095-0.6828) | 67 | (39.0%) | 851 | (12.9%) | 80 | (4.8%) | 7 | (0.8%) |

\*Abbreviations used: Pv: *P. vivax*; FMM: Finite mixture model; Negpop: Negative population model; SL: Superlearner prediction model using different sets of covariates (including the 8 Pf antigens) for predicting recent and historical malaria exposure.

**Supplementary Table 3.**
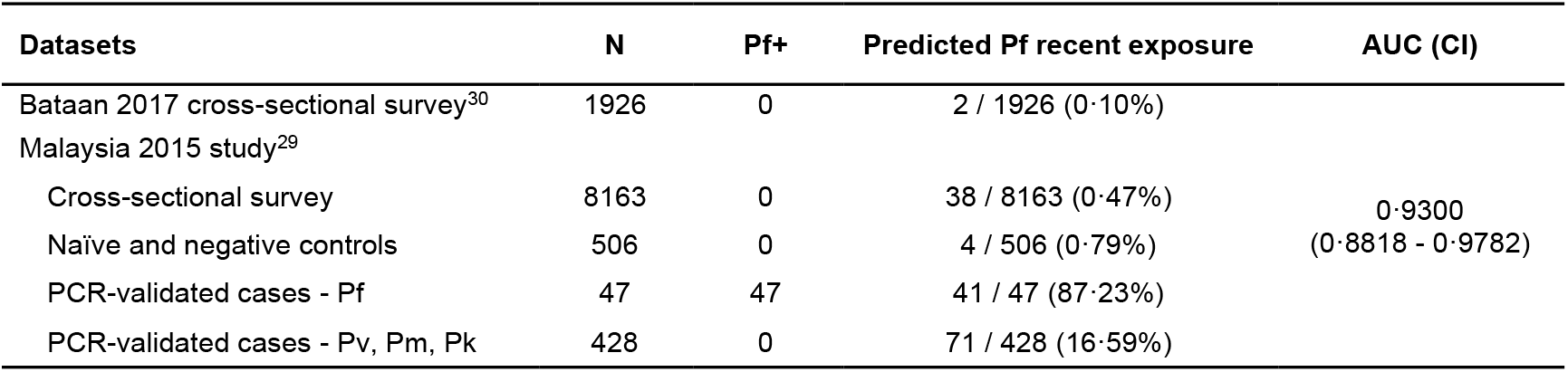
Collated dataset for external validation of RF 4-covariate model for predicting Pf recent exposure

**Supplementary Figure 1.**
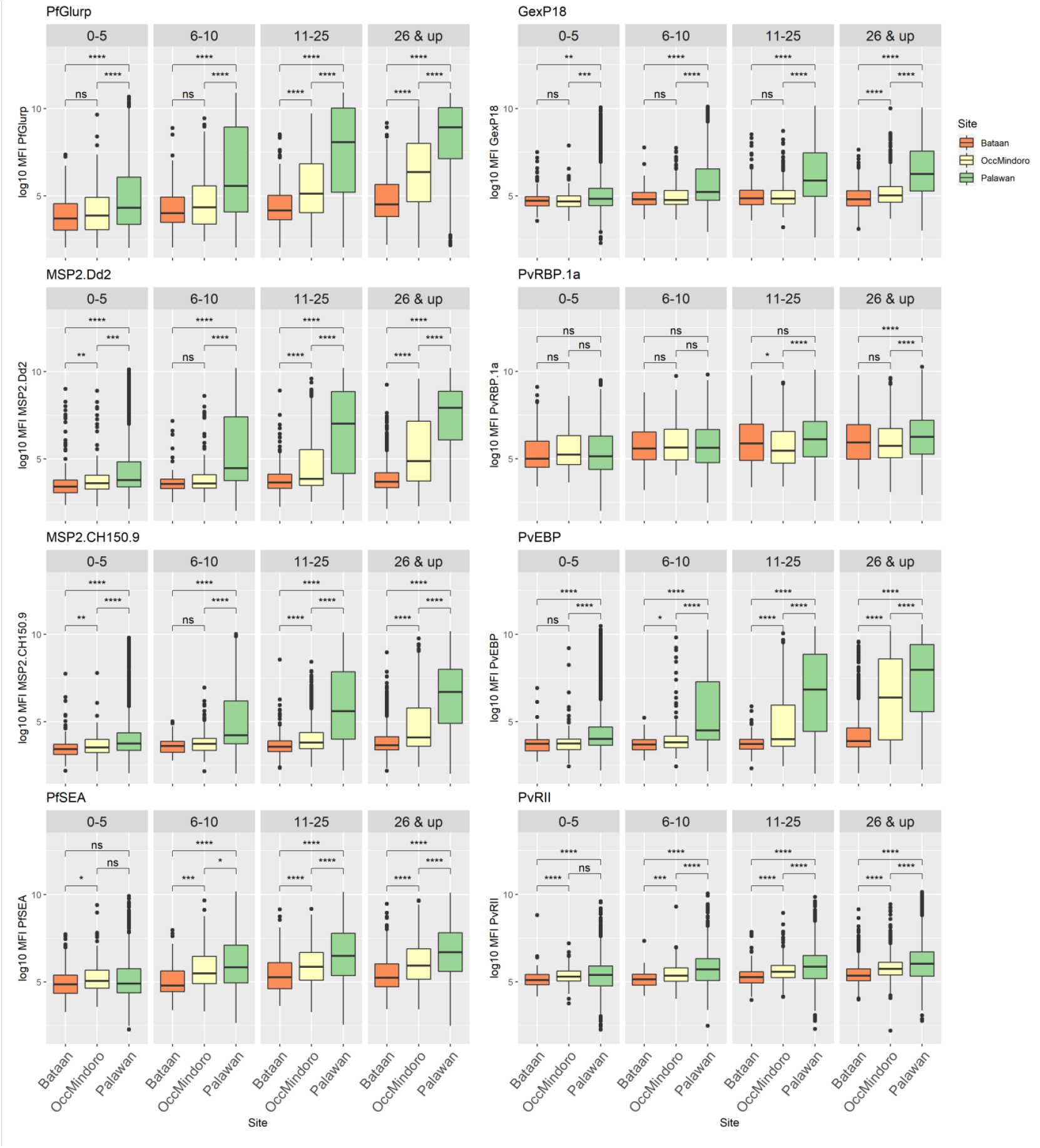
Antibody levels in response to the rest of the *P. falciparum* and *P. vivax* serological markers in the panel, by study site and age group. Statistical difference of overall antibody responses among study sites within age groups were determined using Kruskall-Wallis test and Wilcoxon test for pairwise comparisons. Related to Fig. 1.

**Supplementary Figure 2.**
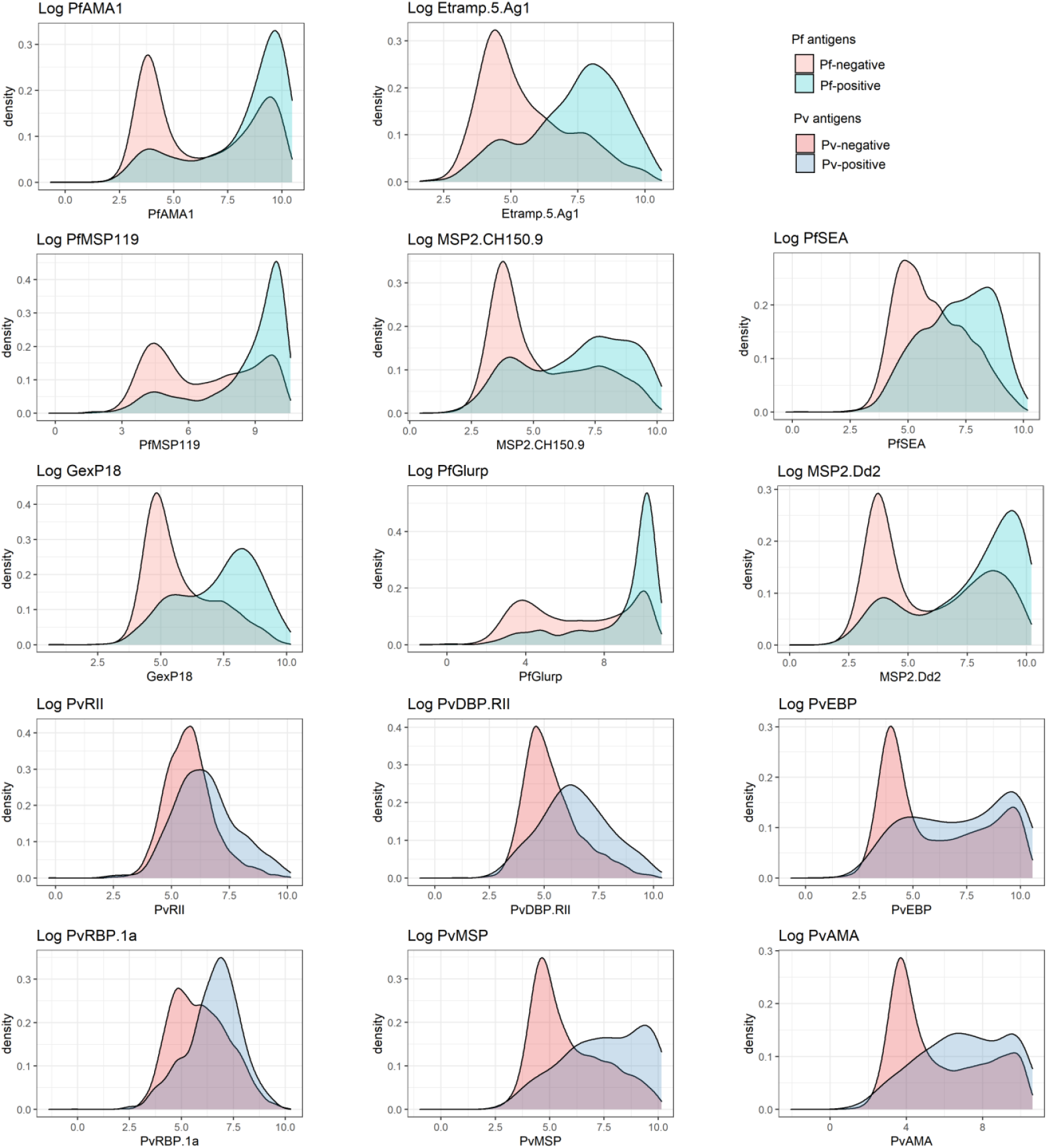
Antibody responses of malaria-negative and malaria-positive populations (presented as density plots of log10 net MFI values) to the Pf and Pv serological markers in the panel (Pf and Pv diagnosed by microscopy, RDT and/or PCR. Related to Fig. 1.

**Supplementary Figure 3.**
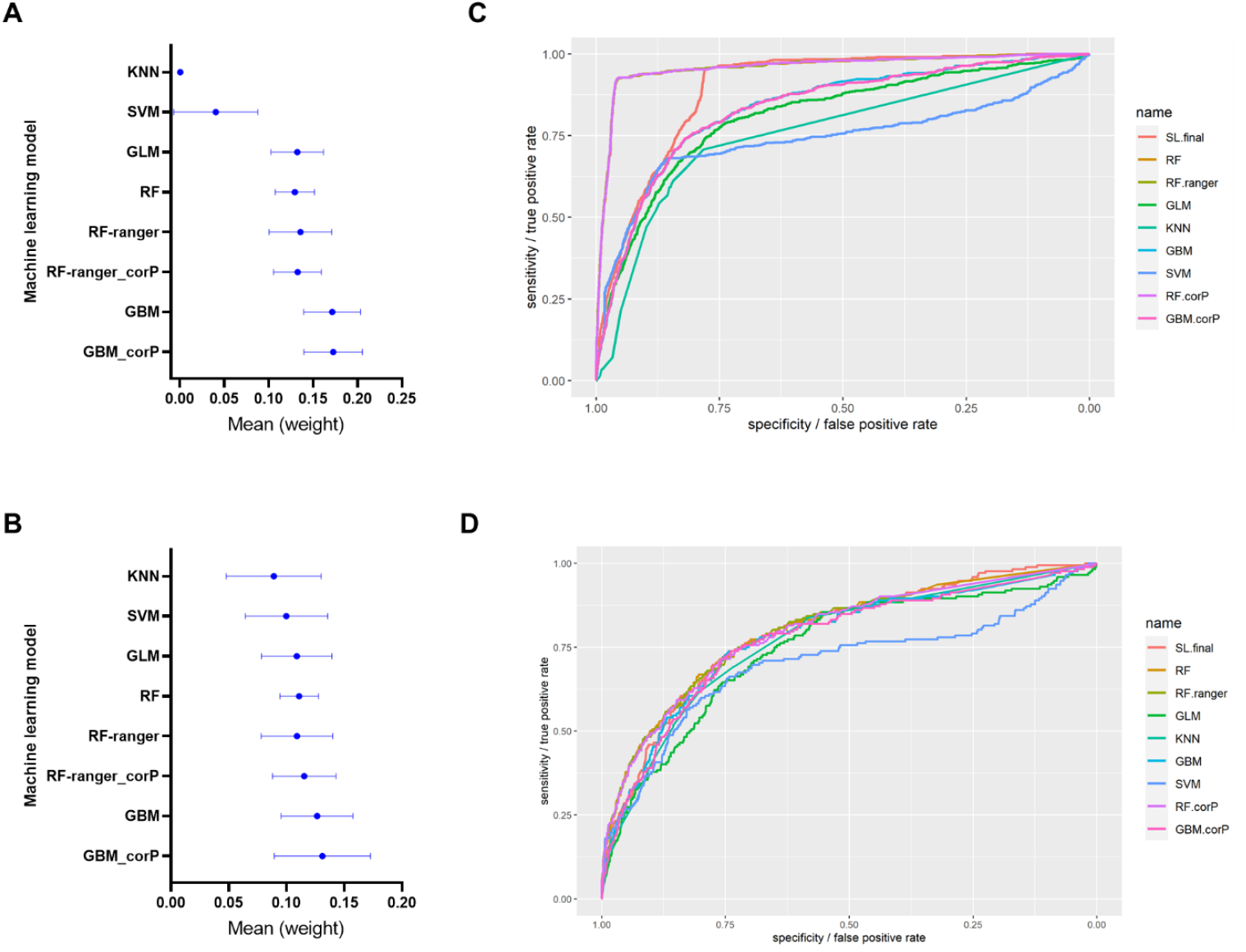
Performances of the individual base learners in the Super Learner ensemble. **A-B)** Plots of the assigned weights to each learner included in the Super Learner models for predicting *P. falciparum* (A) and *P. vivax* recent exposure (B), obtained after 20-fold nested cross-validation of the Pf SL-covar8 and Pv SL-covar6 models. **C-D)** Receiver operating characteristic (ROC) curves for detecting current Pf (C) and Pv (D) infections in the test dataset (n=9132) using the 8 Pf and 6 Pv antigens as covariates, respectively. (SL.final: final Super Learner model, RF: random Forest, RF.ranger: RF from ranger package, kNN: k-Nearest Neighbor, GBM: generalized boosted models (implementation for BRT: boosted regression trees), SVM:Support Vector Machine, and GLM: Generalized linear models; variation of algorithms with “corP” denotes a feature selection that screens for univariate correlation)

## References

1 World Health Organization. Progress towards subnational elimination in the Philippines. Geneva PP - Geneva: World Health Organization, 2014 https://apps.who.int/iris/handle/10665/149678.

2 World Health Organization. World Malaria Report 2019. Geneva, Switzerland, 2019 https://www.who.int/publications-detail/world-malaria-report-2019.

3 Drakeley CJ, Corran PH, Coleman PG, et al. Estimating medium- and long-term trends in malaria transmission by using serological markers of malaria exposure. Proc Natl Acad Sci 2005; 102: 5108–13.

4 malERA Refresh Consultative Panel. malERA: An updated research agenda for characterising the reservoir and measuring transmission in malaria elimination and eradication. PLOS Med 2017; 14: e1002452.

5 World Health Organization, Global Malaria Programme. A Framework for Malaria Elimination. 2017 DOI:Licence: CC BY-NC-SA 3.0 IGO.

6 Cotter C, Sturrock HJW, Hsiang MS, et al. The changing epidemiology of malaria elimination: New strategies for new challenges. Lancet 2013; 382: 900–11.

7 Folegatti PM, Siqueira AM, Monteiro WM, Lacerda MVG, Drakeley CJ, Braga ÉM. A systematic review on malaria sero-epidemiology studies in the Brazilian Amazon: insights into immunological markers for exposure and protection. Malar J 2017; 16: 107.

8 Fowkes FJI, Richards JS, Simpson JA, Beeson JG. The relationship between anti-merozoite antibodies and incidence of Plasmodium falciparum malaria: A systematic review and meta-analysis. PLoS Med 2010; 7. DOI:10.1371/journal.pmed.1000218.

9 Pothin E, Ferguson NM, Drakeley CJ, Ghani AC. Estimating malaria transmission intensity from Plasmodium falciparum serological data using antibody density models. Malar J 2016; 15: 79.

10 Idris ZM, Chan CW, Mohammed M, et al. Serological measures to assess the efficacy of malaria control programme on Ambae Island, Vanuatu. Parasites and Vectors 2017; 10. DOI:10.1186/s13071-017-2139-z.

11 Ssewanyana I, Arinaitwe E, Nankabirwa JI, et al. Avidity of anti - malarial antibodies inversely related to transmission intensity at three sites in Uganda. Malar J 2017; : 1–8.

12 Kerkhof K, Sluydts V, Willen L, et al. Serological markers to measure recent changes in malaria at population level in Cambodia. Malar J 2016; 15: 1–18.

13 van den Hoogen LL, Griffin JT, Cook J, et al. Serology describes a profile of declining malaria transmission in Farafenni, The Gambia. Malar J 2015; 14: 416.

14 Wu L, Mwesigwa J, Affara M, et al. Sero-epidemiological evaluation of malaria transmission in The Gambia before and after mass drug administration. BMC Med 2020; 18: 1–14.

15 Niass O, Saint-Pierre P, Niang M, et al. Modelling dynamic change of malaria transmission in holoendemic setting (Dielmo, Senegal) using longitudinal measures of antibody prevalence to Plasmodium falciparum crude schizonts extract. Malar J 2017; 16: 1–12.

16 Corran P, Coleman P, Riley E, Drakeley C. Serology: a robust indicator of malaria transmission intensity? Trends Parasitol 2007; 23: 575–82.

17 Stanisic DI, Fowkes FJI, Koinari M, et al. Acquisition of antibodies against Plasmodium falciparum merozoites and malaria immunity in young children and the influence of age, force of infection, and magnitude of response. Infect Immun 2015; 83. DOI:10.1128/IAI.02398-14.

18 Fouda GG, Leke RFG, Long C, et al. Multiplex assay for simultaneous measurement of antibodies to multiple Plasmodium falciparum antigens. Clin Vaccine Immunol 2006; 13: 1307–13.

19 Wu L, Hall T, Ssewanyana I, et al. Optimisation and standardisation of a multiplex immunoassay of diverse Plasmodium falciparum antigens to assess changes in malaria transmission using seroepidemiology. Wellcome Open Res 2019; 4: 26.

20 Koffi D, Touré AO, Varela ML, et al. Analysis of antibody profiles in symptomatic malaria in three sentinel sites of Ivory Coast by using multiplex, fluorescent, magnetic, bead-based serological assay (MAGPIX™). Malar J 2015; 14. DOI:10.1186/s12936-015-1043-2.

21 Perraut R, Richard V, Varela ML, et al. Comparative analysis of IgG responses to Plasmodium falciparum MSP1p19 and PF13-DBL1α1 using ELISA and a magnetic bead-based duplex assay (MAGPIX®-Luminex) in a Senegalese meso-endemic community. Malar J 2014; 13. DOI:10.1186/1475-2875-13-410.

22 Ondigo BN, Park GS, Gose SO, et al. Standardization and validation of a cytometric bead assay to assess antibodies to multiple Plasmodium falciparum recombinant antigens. Malar J 2012; 11: 1–17.

23 Helb DA, Tetteh KKA, Felgner PL, et al. Novel serologic biomarkers provide accurate estimates of recent Plasmodium falciparum exposure for individuals and communities. Proc Natl Acad Sci 2015; 112: E4438–47.

24 Wu L, Mwesigwa J, Affara M, et al. Antibody responses to a suite of novel serological markers for malaria surveillance demonstrate strong correlation with clinical and parasitological infection across seasons and transmission settings in The Gambia. BMC Med 2020; : 2020.07.10.20067488.

25 Longley RJ, White MT, Takashima E, et al. Development and validation of serological markers for detecting recent Plasmodium vivax infection. Nat Med 2020; 26: 741–9.

26 Reyes RA, Fornace KM, Macalinao MLM, et al. Enhanced health facility surveys to support malaria control and elimination across different transmission settings in the Philippines. Am J Trop Med Hyg 2021; 104: 968–78.

27 Coutts SP, King JD, Pa’au M, et al. Prevalence and risk factors associated with lymphatic filariasis in American Samoa after mass drug administration. Trop Med Health 2017; 45: 1–10.

28 van den Hoogen LL, Présumé J, Romilus I, et al. Quality control of multiplex antibody detection in samples from large-scale surveys: the example of malaria in Haiti. Sci Rep 2020; 10: 1–10.

29 Fornace KM, Brock PM, Abidin TR, et al. Environmental risk factors and exposure to the zoonotic malaria parasite Plasmodium knowlesi across northern Sabah, Malaysia: a population-based cross-sectional survey. Lancet Planet Heal 2019; 3: e179–86.

30 van den Hoogen LL, Bareng P, Alves J, et al. Comparison of Commercial ELISA Kits to Confirm the Absence of Transmission in Malaria Elimination Settings. Front Public Heal 2020; 8: 1–12.

31 Hubbard A, Munoz ID, Decker A, et al. Time-dependent prediction and evaluation of variable importance using superlearning in high-dimensional clinical data. J Trauma Acute Care Surg 2013; 75: 1–12.

32 Arnold BF, van der Laan MJ, Hubbard AE, et al. Measuring changes in transmission of neglected tropical diseases, malaria, and enteric pathogens from quantitative antibody levels. PLoS Negl Trop Dis 2017; 11: 1–20.

33 Sepúlveda N, Stresman G, White MT, Drakeley CJ. Current mathematical models for analyzing antimalarial antibody data with an eye to malaria elimination and eradication. J Immunol Res 2015; 2015. DOI:10.1155/2015/738030.

34 Fornace KM, Surendra H, Abidin TR, et al. Use of mobile technology-based participatory mapping approaches to geolocate health facility attendees for disease surveillance in low resource settings. Int J Health Geogr 2018; 17: 21.

35 Fornace KM, Herman LS, Abidin TR, et al. Exposure and infection to Plasmodium knowlesi in case study communities in Northern Sabah, Malaysia and Palawan, The Philippines. PLoS Negl Trop Dis 2018; 12: e0006432.

36 van den Hoogen LL, Stresman G, Présumé J et al. Selection of Antibody Responses Associated With Plasmodium falciparum Infections in the Context of Malaria Elimination. Front Immunol 2020; 11: 1–12.

37 Rogier E, Nace D, Dimbu PR, et al. Framework for Characterizing Longitudinal Antibody Response in Children After Plasmodium falciparum Infection. Front Immunol 2021; 12: 1–11.

38 Chotirat S, Nekkab N, Kumpitak C, et al. Application of 23 novel serological markers for identifying recent exposure to <em>Plasmodium vivax</em> parasites in an endemic population of western Thailand. medRxiv 2021; : 2021.03.01.21252492.

39 Rosado J, Pelleau S, Cockram C, et al. Multiplex assays for the identification of serological signatures of SARS-CoV-2 infection: an antibody-based diagnostic and machine learning study. The Lancet Microbe 2020; 5247: 1–10.

40 Biggs J, Raman J, Cook J, et al. Serology reveals heterogeneity of Plasmodium falciparum transmission in northeastern South Africa: Implications for malaria elimination. Malar J 2017; 16. DOI:10.1186/s12936-017-1701-7.

41 Rosas-Aguirre A, Speybroeck N, Llanos-Cuentas A, et al. Hotspots of malaria transmission in the Peruvian amazon: Rapid assessment through a parasitological and serological survey. PLoS One 2015; 10. DOI:10.1371/journal.pone.0137458.

42 World Health Organization. WHO malaria policy advisory committee meeting: meeting report, October 2017. Geneva, Switzerland, 2017 https://www.who.int/malaria/mpac/mpac-oct2017-erg-malaria-low-density-infections-session2.pdf?ua=1.

43 Fowkes FJI, Boeuf P, Beeson JG. Immunity to malaria in an era of declining malaria transmission. Parasitology. 2016; 143: 139–53.

44 Collins CR, Withers-Martinez C, Bentley GA, Batchelor AH, Thomas AW, Blackman MJ. Fine mapping of an epitope recognized by an invasion-inhibitory monoclonal antibody on the malaria vaccine candidate apical membrane antigen 1. J Biol Chem 2007; 282: 7431–41.

45 Burghaus PA, Holder AA. Expression of the 19-kilodalton carboxy-terminal fragment of the Plasmodium falciparum merozoite surface protein-1 in Escherichia coli as a correctly folded protein. Mol Biochem Parasitol 1994; 64: 165–9.

46 Theisen M, Vuust J, Gottschau A, Jepsen S, Hogh B. Antigenicity and immunogenicity of recombinant glutamate-rich protein of Plasmodium falciparum expressed in Escherichia coli. Clin Diagn Lab Immunol 1995; 2: 30–4.

47 Spielmann T, Fergusen DJP, Beck H-P. etramps, a New Plasmodium falciparum Gene Family Coding for Developmentally Regulated and Highly Charged Membrane Proteins Located at the Parasite–Host Cell Interface. Mol Biol Cell 2003; 14: 1529–44.

48 van den Hoogen LL, Walk J, Oulton T, et al. Antibody Responses to Antigenic Targets of Recent Exposure Are Associated With Low-Density Parasitemia in Controlled Human Plasmodium falciparum Infections. Front Microbiol 2019; 9: 1–11.

49 Raj DK, Nixon CP, Nixon CE, et al. Antibodies to PfSEA-1 block parasite egress from RBCs and protect against malaria infection. Science (80-) 2014; 344: 871–7.

50 Polley SD, Conway DJ, Cavanagh DR, et al. High levels of serum antibodies to merozoite surface protein 2 of Plasmodium falciparum are associated with reduced risk of clinical malaria in coastal Kenya. Vaccine 2006; 24: 4233–46.

51 Taylor RR, Smith DB, Robinson VJ, McBride JS, Riley EM. Human antibody response to Plasmodium falciparum merozoite surface protein 2 is serogroup specific and predominantly of the immunoglobulin G3 subclass. Infect Immun 1995; 63: 4382–8.

52 Chuquiyauri R, Molina DM, Moss EL, et al. Genome-scale protein microarray comparison of human antibody responses in plasmodium vivax relapse and reinfection. Am J Trop Med Hyg 2015; 93: 801–9.

53 França CT, Hostetler JB, Sharma S, et al. An Antibody Screen of a Plasmodium vivax Antigen Library Identifies Novel Merozoite Proteins Associated with Clinical Protection. PLoS Negl Trop Dis 2016; 10: 1–15.

54 França CT, He WQ, Gruszczyk J, et al. Plasmodium vivax Reticulocyte Binding Proteins Are Key Targets of Naturally Acquired Immunity in Young Papua New Guinean Children. PLoS Negl Trop Dis 2016; 10: 1–17.

55 Ntumngia FB, Schloegel J, Barnes SJ, et al. Conserved and variant epitopes of Plasmodium vivax duffy binding protein as targets of inhibitory monoclonal antibodies. Infect Immun 2012; 80: 1203–8.

56 Hester J, Chan ER, Menard D, et al. De Novo Assembly of a Field Isolate Genome Reveals Novel Plasmodium vivax Erythrocyte Invasion Genes. PLoS Negl Trop Dis 2013; 7. DOI:10.1371/journal.pntd.0002569.

57 Menard D, Chan ER, Benedet C, et al. Whole Genome Sequencing of Field Isolates Reveals a Common Duplication of the Duffy Binding Protein Gene in Malagasy Plasmodium vivax Strains. PLoS Negl Trop Dis 2013; 7. DOI:10.1371/journal.pntd.0002489.

